# Moderate-to-vigorous Intensity Physical Activity and Incident Left-sided Degenerative Valvular Heart Disease

**DOI:** 10.1101/2023.08.21.23294391

**Authors:** Ziang Li, Sijing Cheng, Bo Guo, Lu Ding, Yu Liang, Yinghan Shen, Jinyue Li, Yiqing Hu, Tianxin Long, Xinli Guo, Junbo Ge, Runlin Gao, Philippe Pibarot, Bin Zhang, Haiyan Xu, Marie-Annick Clavel, Yongjian Wu

## Abstract

**Background:** Despite the escalating incidence of degenerative valvular heart disease (VHD), recommended preventive interventions are conspicuously absent. Physical activity has proven effective in preventing atherosclerotic cardiovascular disease, but its role in preventing VHD remains uncertain. This study aimed to explore the association between moderate-to-vigorous intensity physical activity (MVPA) and incident left-sided degenerative VHD in middle-aged adults from the UK biobank.

**Methods:** Data from wrist-worn accelerometer and self-reported questionnaires were utilized to assess the impact of MVPA volume on the incidence of aortic valve stenosis (AS), aortic valve regurgitation (AR), and mitral valve regurgitation (MR). Incident VHD were ascertained from hospital admissions and death reports. Cox proportional hazards regression models were employed to calculate hazard ratios (HRs) and 95% confidence intervals (CIs), adjusted for sociodemographic characteristics, lifestyle risk factors, and comorbidities.

**Results:** In the accelerometer-derived MVPA cohort (n=90,865; median age 63; 43% male; median follow-up 8.1 years), the age- and sex-adjusted incidence rates per 1000 person-years were 0.70 for AS, 0.29 for AR, and 0.84 for MR. In the questionnaire-based MVPA cohort (n=397,335; median age 57; 47% male; median follow-up 13.8 years), the corresponding rates were 0.76 for AS, 0.29 for AR, and 0.76 for MR. The accelerometer-measured MVPA volume showed a curvilinear relationship with reduced risk of AS, plateauing above 300 min/week. Participants engaging in 150-299 minutes of MVPA per week, meeting the guideline recommendation, had the most significant reduction in AS risk compared to those with no MVPA [adjusted HR, 0.53 (95% CI, 0.37-0.76)]. Similar results were found in the questionnaire-based MVPA cohort, with 150-299 minutes of MVPA showing a relatively smaller reduction in HR [adjusted HR, 0.82 (95% CI, 0.73-0.91)]. The association remained consistent across subgroups at high risk for AS. However, there was no significant inverse association of MVPA with risk of AR or MR.

**Conclusion:** Adhering to the recommended MVPA duration (150-299 min/week) was associated with the lowest risk of developing AS. Encouraging the utilization of wearable devices to monitor activity levels enhances AS risk reduction. Nonetheless, MVPA’s efficacy in preventing valvular regurgitation is limited, revealing distinctive pathological mechanisms in valvular stenotic and regurgitation lesions.

**Clinical Perspective:** *What Is New?:*

- Engaging in 150-299 minutes of moderate-to-vigorous intensity physical activity per week can reduce the risk of aortic valve stenosis by nearly 50% in middle-aged individuals.
- Using wearable devices to measure activity levels may improve the risk stratification of aortic valve stenosis compared to assessments based on questionnaires.
- No significant association is observed between moderate-to-vigorous intensity physical activity and risks of aortic valve regurgitation and mitral valve regurgitation.

*What Are the Clinical Implications?:*

- Objective activity monitoring through wearable devices shows promise as an effective nonpharmaceutical intervention to alleviate the healthcare burdens associated with aortic valve stenosis.
- Encouraging middle-aged individuals at higher risk for aortic valve stenosis to engage in moderate volume (150-299 minutes per week) and moderate intensity physical activity (e.g., walking at a speed of 2.5 miles per hour) is recommended.
- Engaging in physical activity beyond the recommended volume and intensity does not yield additional benefits nor pose additional risks for aortic valve stenosis.

## Introduction

Degenerative valvular heart disease (VHD) is the predominant form of valvular heart lesions in Western countries, primarily occurring in individuals in their fifth decade and beyond, with left-sided valve involvement being the most prevailing manifestation.^1^ The aging population has contributed to a rapid increase in the incidence and prevalence of degenerative VHD, projected to double by 2050 among the elderly.^2, 3^ Despite the growing epidemic trend and significant disease burden, there are currently no recommended preventive measures effectively thwarting VHD.^4^

Compelling evidence supports the role of physical activity in the prevention of atherosclerotic cardiovascular disease and related mortality.^5, 6^ A recent meta-analysis involving over 30 million participants from 94 prospective cohorts found that adhering to the World Health Organization (WHO) recommendation of 150 minutes per week of moderate-to-vigorous intensity physical activity (MVPA) was associated with a 21% lower risk of coronary heart disease.^5^ Considering the favorable effects of physical activity on cardiorespiratory fitness, metabolic milieu, and cardiac tissue adaptations, it is reasonable to expect its beneficial impact on preventing valvular degeneration as well.^7, 8^ However, the existing evidence has not provided a clear understanding of the association between physical activity and the risk of VHD. Recent epidemiological studies showed that questionnaire-based physical activity did not decrease the risk of aortic valve stenosis (AS).^9, 10^ These studies, characterized by small sample sizes, non-standardized questionnaire content, and reliance on self-reported assessments, are susceptible to measurement error and recall bias, limiting the exploration of the impact of physical activity on the natural history of degenerative VHD. Additionally, the impact of vigorous intensity physical activity (VPA) on valve function remains poorly understood, with some hypotheses proposing that the increased hemodynamic loads during exercise may contribute to the elevated risk of VHD.^11, 12^ Therefore, a deficiency exists in large-scale prospective studies that investigate the effects of physical activity volume, intensity, and their joint influence on the natural progression of VHD.

The WHO Physical Activity Guidelines Panel has recently recommended using wearable devices to assess the relationship between physical activity and disease risk.^13^ Wearable devices possess objective measurement capabilities and continuous monitoring ability over a prolonged period, making them optimal tools for quantifying and tracking activity levels. Moreover, consumer wearable devices offer a user-friendly way to self-monitor activity levels, with their utilization expected to increase by 25% annually until 2025.^14^ Thus, in the present study, we utilized accelerometer-measured and self-reported physical activity data from the UK Biobank to investigate the dose-response association between MVPA and the risks of left-sided degenerative VHD.

## Methods

### Data sources

The UK Biobank is a prospective cohort study comprising more than 500,000 adults aged 37–73 years recruited from the general population across the United Kingdom between 2006 and 2010.^15^ Ethical approval for the UK Biobank was granted by the North West Multicenter Research Ethics Committee (reference number: 11/NW/0382), and all participants provided written informed consent. This study was conducted based on the UK Biobank cohort study under application number 91305. The reporting of this study followed the “Strengthening the Reporting of Observational Studies in Epidemiology” (STROBE) reporting guideline (Table S1).^16^

### Accelerometer-measured physical activity

From 2013 to 2015, a sub-sample of over 100,000 randomly selected participants from the UK Biobank wore accelerometers on their dominant wrist to measure their weekly physical activity volume.^17^ The intensity of physical activity was quantified by assessing the average vector magnitude in 5-second epochs, measured in milligravity (mg) units. This approach has been validated through comparison with doubly labeled water method for estimating energy expenditure.^18^ The average vector magnitude exceeding 100 mg serves as the threshold for determining moderate-to-vigorous intensity activity.^19, 20^ Previous studies computed the volume of MVPA in bouts, defined as 5-minute periods where more than 80% of 5-second epochs had an average vector magnitude ≥100 mg.^21-23^ However, considering that participants’ activity durations may not align with five-minute increments, we have optimized the approach by precisely calculating the “remnant MVPA volume”. This strategy captures activity occurring at the end of bouts, lasting less than 5 minutes, identified by 80% of the 5-second epochs within each minute having an average vector magnitude ≥100 mg. This refinement enables a more accurate estimation of weekly physical activity duration for each participant, resulting in an increase in median MVPA from 140 min/week [interquartile range (IQR): 60-255] to 166 min/week [IQR: 73-300] (Figure S1).

For participants who provided less than 7 days of accelerometry data (median wear duration 6.93, IQR [6.73, 7.00]), the MVPA volume was extrapolated to a one-week duration, considering that physical activity thresholds are typically defined on a weekly basis.^23^ According to the current guidelines recommending 150-299 minutes of MVPA per week for adults, we categorized the participants into four groups based on their weekly MVPA duration: 0 minutes, 1-149 minutes, 150-299 minutes, and ≥300 minutes.^24, 25^ ^26^ Additionally, the volumes of VPA and moderate-intensity physical activity (MPA) were calculated for each participant based on their total MVPA time, with VPA determined by assessing the average vector magnitude exceeding 400 mg.^23, 27^

### Questionnaire-based physical activity

Between 2006 and 2010, sociodemographic information, lifestyle data, and health assessments, including self-reported physical activity data, were collected from over 400,000 participants in the UK Biobank study. Self-reported physical activity data were obtained using the short-version of the International Physical Activity Questionnaire (IPAQ), which assesses participants’ weekly duration of engagement in three activity types: light-intensity, moderate-intensity, and vigorous-intensity activities.^28, 29^ The total volume of MVPA was computed by summing the durations of moderate-intensity and vigorous-intensity activities. Similar to the categorization based on accelerometer-measured physical activity, we classified participants into four groups based on their self-reported weekly MVPA volume: 0 minutes, 1-149 minutes, 150-299 minutes, and ≥300 minutes.

### Study cohorts

Two observational cohorts were formed based on distinct modalities of physical activity assessment: objective evaluation using wrist-worn accelerometers and subjective evaluation through self-reported questionnaires. The primary cohort included 90,865 participants who had valid accelerometer-measured physical activity data and did not have prevalent VHD or heart failure (Figure 1). The secondary cohort comprised 397,335 participants who met the same exclusion criteria and completed a self-reported physical activity questionnaire at recruitment. Detailed exclusion criteria and corresponding ICD-10 codes were presented in Figure S2 and Table S2, respectively. All participants in the primary cohort were followed from the start of accelerometer wear, while in the secondary cohort, they were followed from the date of participant consent to join the UK Biobank. The follow-up period continued until the incidence of left-sided degenerative VHD, death, or the end of the study period (December 31, 2022), whichever came first.

**Figure 1.**
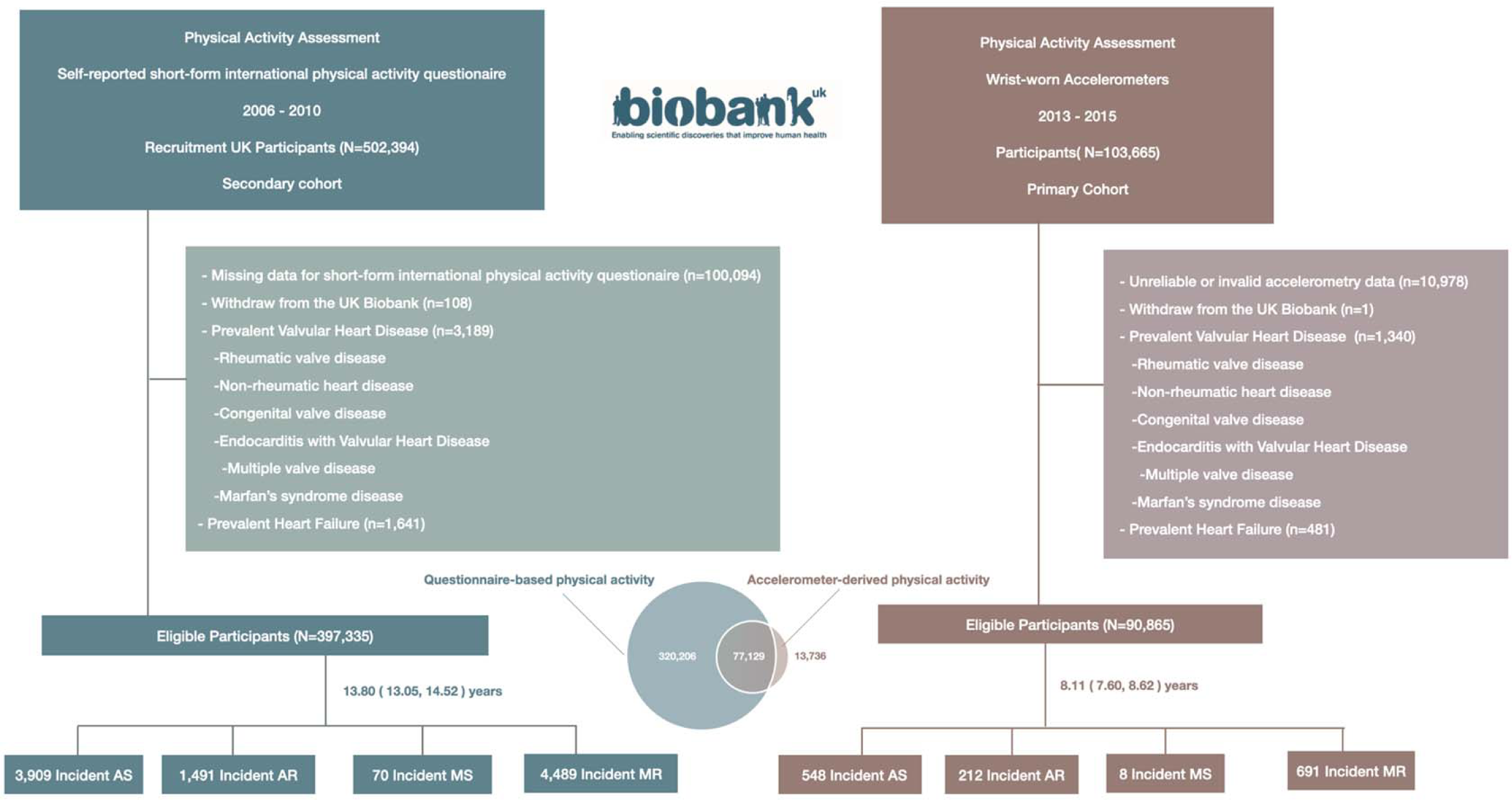
Flowchart of the study. Venn diagram illustrates the number of individuals within the population who engaged in accelerometer-measured physical activity and questionnaire-based physical activity. AS, aortic valve stenosis; AR, aortic regurgitation; MS: mitral valve stenosis; MR: mitral valve regurgitation.

### Assessment of VHD

We extracted specific incident events related to AS (ICD-10 codes, I35.0), aortic valve regurgitation (AR) (ICD-10 codes, I35.1), mitral valve stenosis (MS) (ICD-10 codes, I34.2), and mitral valve regurgitation (MR) (ICD-10 codes, I34.0) from hospital episode records and death registers.^30^ To ensure the absence of prevalent valvular heart disease in the study cohorts, individuals with a documented diagnosis of valvular heart disease at baseline, including rheumatic valve disease, degenerative valve disease (also known as non-rheumatic VHD), congenital valve disease, endocarditis with valvular heart disease, and Marfan’s syndrome, were excluded based on records obtained from self-report, primary care, or hospital admission sources (Figure S2 & Table S2).

### Covariates

Age was determined based on the date of birth of each participant and the recruitment time of cohorts. Socio-demographic information, including sex (male or female), ethnicity (white or non-white), Townsend deprivation index, and educational attainment (university degree or not), as well as comprehensive lifestyle factors such as smoking status (never, previous, or current), alcohol consumption, diet score, sleep score, and discretionary screen-time, were collected through standardized touchscreen questionnaires. Alcohol consumption (g/day) was estimated based on the frequency of alcohol intake and the standardized amounts of various beverage types. The standardized alcohol units (1unit=8g) in different beverage categories were sourced from https://www.drinkaware.co.uk/. Detailed information was provided in Table S3.

The Diet score assessed participants’ adherence to regular dietary intake, including the consumption of fruits, vegetables, fish, red meat, and processed meat (Table S4). ^28, 31^ The sleep score evaluated participants’ sleep quality based on five sleep indices: morning chronotype, sleep duration, insomnia, snoring, and daytime sleepiness (Table S5).^32, 33^ Both the diet score and sleep score range from 0 to 3 and 0 to 5, respectively, with higher scores indicating a healthier pattern. Discretionary screen-time was defined as the total daily hours spent on watching TV and non-occupational computer use. Clinical comorbidities, including hypertension, obesity, type 2 diabetes, dyslipidemia, ischemic heart disease, stroke, cardiomyopathy, atrial fibrillation, chronic obstructive pulmonary disease, and end-stage renal disease, were identified based on self-report, primary care, or hospital admission records at baseline. Prevalent cancer diagnoses were identified through linkage with national cancer registries.

### Statistical analysis

Descriptive characteristics by categories of MVPA were summarized as mean (standard deviation), median [IQR], or percentage as appropriate (Table 1). The distributions of physical activity volume, as derived by accelerometers or questionnaires, were presented using histograms stratified by gender. Age- and gender-adjusted incidence rates of left-sided degenerative VHD within MVPA volume groups were estimated using Poisson regression and reported as the number of events per 1,000 person-years. Missing covariate data in the two cohorts, ranging from 0.01% to 1.83% for each variable, were addressed using multiple imputation by chained equations (Table S6). The imputed overall samples, serving as the main study population, had similar baseline characteristics to the complete samples, which excluded any missing covariate data from the overall samples (Table S7).

**Table 1.**
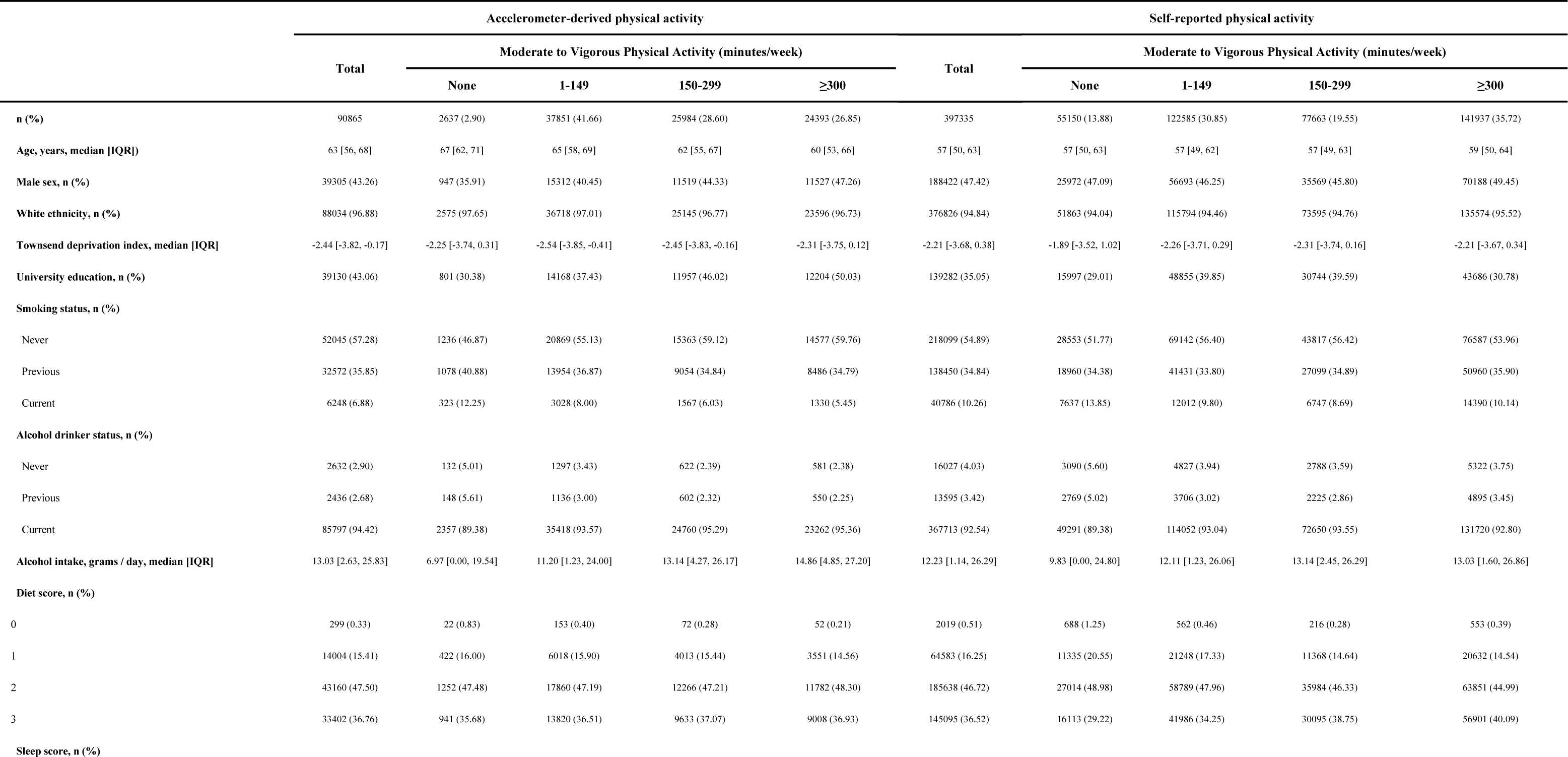
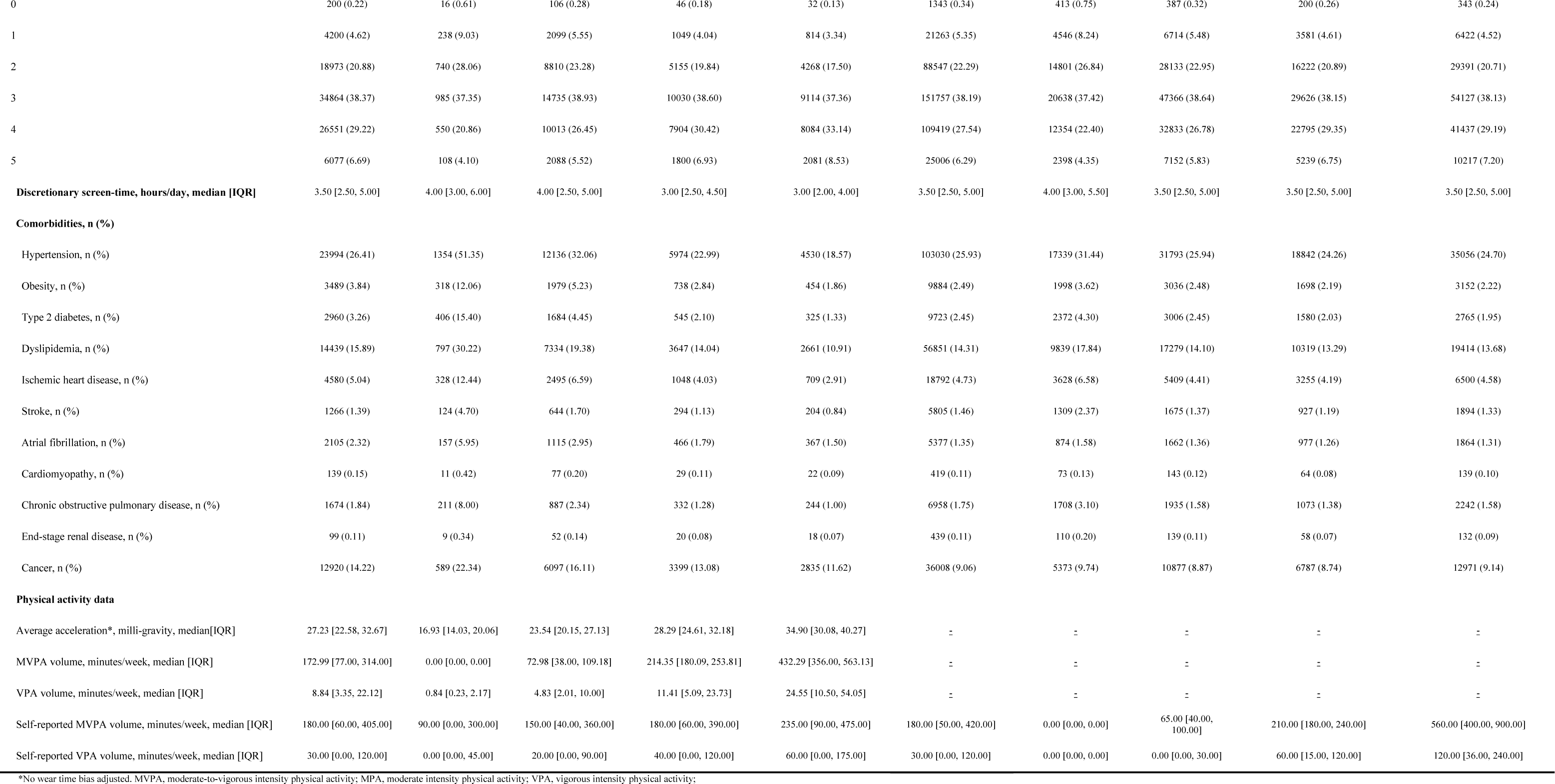
Participants descriptive characteristics by guideline-recommended moderate-to-vigorous physical activity levels.

To evaluate the potential non-linear association between MVPA volume and incident left-sided degenerative VHD, restricted cubic splines were initially employed. The reference point was set as 0 min/week, equivalent to the 3rd percentile of the accelerometer-derived MVPA volume distribution and the 14th percentile of the questionnaire-based MVPA volume distribution. For categorical analyses, MVPA was classified based on adherence to recommended weekly physical activity duration as well as quartiles and deciles of the MVPA volume distribution. Cumulative risk curves were generated to illustrate the standardized risks of mortality outcomes across different MVPA volume groups. Cox proportional hazard analyses were conducted to estimate hazard ratios (HRs) along with 95% confidence intervals (CIs). The proportional hazards assumption was verified by the residual plot. According to a priori defined directed acyclic graph (Figure S3), all the aforementioned analyses were adjusted for the following covariates: age, sex, ethnicity, Townsend deprivation index, educational attainment, smoking status, alcohol consumption, diet score, sleep score, discretionary screen-time, and clinical comorbidities, including hypertension, obesity, type 2 diabetes, dyslipidemia, ischemic heart disease, stroke, cardiomyopathy, atrial fibrillation, chronic obstructive pulmonary disease, end-stage renal disease, and cancer.

In addition, a series of sensitivity analyses were performed. Firstly, we employed the Fine and Gray competing risks regression model to address the competing risk associated with death from other causes. Secondly, to minimize the potential bias of reverse causality, individuals who experienced an incident degenerative VHD event within the initial 2 years of follow-up were excluded from the analysis.^20, 34^ Thirdly, analyses were restricted to participants with complete covariate data. Fourth, repeated Cox analyses were conducted by substituting the binary covariates of hypertension, obesity, dyslipidemia, diabetes and end-stage renal disease with the continuous variables of systolic blood pressure, body mass index, lipoprotein(a), low-density lipoprotein cholesterol, glycated hemoglobin A1c and estimated glomerular filtration rate, all of which are established causal factors for degenerative VHD.

Subgroup analyses were performed to investigate the relationship within potential effect modifiers. Multiplicative interaction analyses were conducted, comparing models with and without a cross-product term using likelihood ratio tests. Moreover, the 5-year absolute risk of AS in the primary cohort and the 10-year absolute risk in the secondary cohort were estimated. Age, sex, and physical activity categories were included as covariates in the Fine and Gray’s competing risks regression model, and various combinations of covariate values were utilized to estimate the absolute risks. We performed an exploratory analysis using mutually adjusted Cox proportional hazards regression models to explore the relationship between MPA and VPA with the incidence of AS. We further examined the joint association of MPA (0 min/week, 1-149 min/week, 150-299 min/week, ≥300 min/week) and VPA (0 min/week, 1-74 min/week, 75-149 min/week, ≥150 min/week). The reference group for this analysis consisted of individuals who had the lowest levels of both MPA (0 min/week) and VPA (0 min/week). All analyses were performed using R version 4.2.2. Bonferroni-corrected significance level of p-value < 0.017 (0.05/3) was used to adjust for multiple testing of three VHD outcomes.

## Result

### Sample characteristics

Among the 90,865 individuals in the primary cohort (median age [IQR]: 63 [56, 68] years; 43% male), a total of 548 events of AS, 212 events of AR, 8 events of MS, and 691 events of MR were observed during a median follow-up of 8.11 years (IQR, 7.60-8.62 years). In the secondary cohort, consisting of 397,335 individuals (median age [IQR]: 57 [50, 63] years; 47% male), a total of 3,909 events of AS, 1,491 events of AR, 70 events of MS, and 4,4981 events of MR were observed during a median follow-up of 13.80 years (IQR, 13.05-14.52 years). Due to the limited number of incident MS events (8 events in the primary cohort; 70 events in the secondary cohort) and insufficient statistical power (less than 10 events per variable), the impact of MVPA on incident MS was not assessed in this study.

Participants with higher MVPA volume, as measured by accelerometers, tended to be younger, more likely to be men, with a higher educational level, less likely to smoke, reported better sleep quality, and had fewer clinical comorbidities (Table 1). However, these trends were not statistically significant in the self-reported MVPA cohort (Table 1). A more compact distribution was observed for the accelerometer-derived MVPA volume compared to the questionnaire-based MVPA volume (Figure 2). The median accelerometer-derived MVPA volume was 173 (IQR, 77-314) minutes per week, with 55.44% of individuals meeting the recommended guidelines. Similarly, the median questionnaire-based MVPA volume was 180 (IQR, 50-420) minutes per week, with 55.27% of individuals meeting the recommended guidelines.

**Figure 2.**
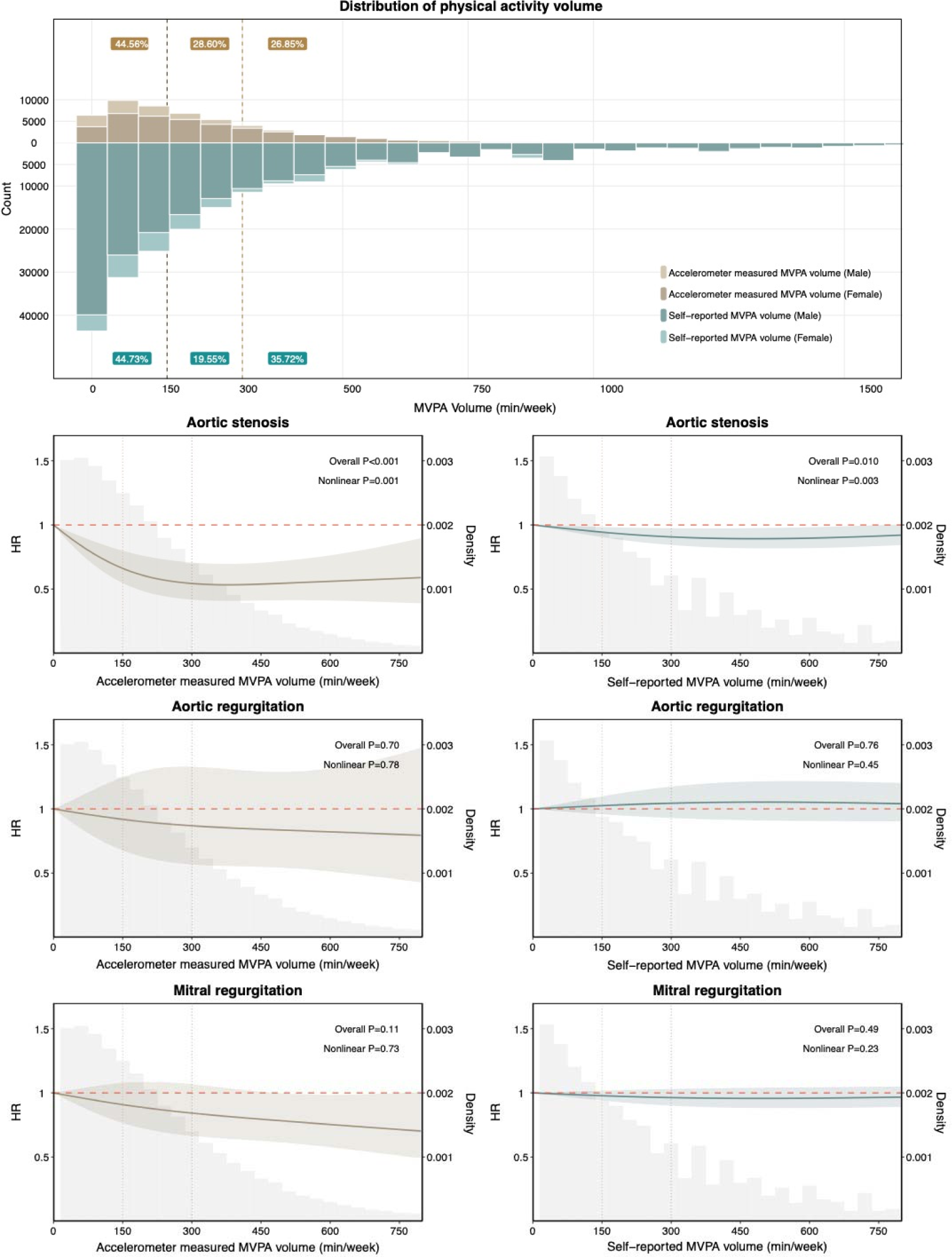
Moderate-to-vigorous intensity physical activity volume distribution and dose-response association with incident left-sided degenerative valvular heart disease. Upper panel. Distribution of moderate-to-vigorous intensity physical activity volume. The brown box represents the percentage of the population according to wrist-worn accelerometer data, while the green box represents the percentage based on questionnaire responses. Lower panel. The left column represents the association between accelerometer-measured physical activity and the incidence of valve heart disease. The right column represents the association between questionnaire-based physical activity and the incidence of valve heart disease. The reference point was set as 0 min/week, equivalent to the 3rd percentile of the accelerometer-derived MVPA volume distribution and the 14th percentile of the questionnaire-based MVPA volume distribution. The gray area indicates the density of the population. Adjusted for age, sex, ethnicity, Townsend deprivation index, educational attainment, smoking status, alcohol consumption, diet score, sleep score, discretionary screen-time, and clinical comorbidities, including hypertension, obesity, type 2 diabetes, dyslipidemia, ischemic heart disease, stroke, cardiomyopathy, atrial fibrillation, chronic obstructive pulmonary disease, end-stage renal disease, and cancer. MVPA, moderate-to-vigorous intensity physical activity.

### Dose-response relationship between MVPA volume and VHD incidence

Both accelerometer-derived and questionnaire-based assessments of MVPA volume exhibited a curvilinear association with decreased AS risk (all nonlinear p<0.004) (Figure 2). The inverse association was more pronounced for accelerometer-derived MVPA volume within the range of 1-300 min/week and reached a plateau beyond 300 min/week, with no additional benefits observed beyond this threshold. While the associations of questionnaire-based MVPA with AS incidence were generally consistent in direction, the effect sizes were smaller compared to accelerometer-derived MVPA. No significant associations were found between MVPA volume and the risk of AR and MR in both cohorts (all p overall >0.05).

### Multivariable-adjusted associations of MVPA volume with VHD incidence

The age- and sex-adjusted incidence rate of AS was 1.96 (1.45, 2.66) per 1000 person-years for participants with 0 min/week of accelerometer-measured MVPA, 0.93 (0.83, 1.04) for those with 1-149 min/week of MVPA, 0.54 (0.45, 0.66) for those with 150-299 min/week of MVPA, and 0.56 (0.45, 0.70) for those with ≥300 min/week of MVPA (Figure 3). In multivariable-adjusted analyses, participants who engaged in accelerometer-measured MVPA volume of 150-299 min/week, as recommended by the guidelines, demonstrated the lowest incidence of AS (hazard ratio [HR], 0.53; 95% confidence interval [CI], 0.37-0.76) compared to individuals with 0 min/week of MVPA (Figure 4). Consistent findings were observed across different MVPA grouping strategies: the third quartile (174-314 min/week) of MVPA exhibited the lowest risk of AS incidence (HR, 0.61; 95% CI, 0.47-0.78) compared to the lowest quartile (0-77 min/week) (Figure 4); Individuals in the sixth decile (173-220 min/week), seventh decile (221-278 min/week), and eighth decile (279-357 min/week) of MVPA demonstrated the top three lowest risk of AS incidence, with reductions of 50%, 49%, and 48%, respectively, compared to the lowest decile (0-26 min/week) of MVPA (Figure S5). Additionally, this finding was similar using questionnaire-based MVPA, although the effect sizes were smaller (Figure 4 & Figure S5). Participants who engaged in questionnaire-based MVPA volume of 150-299 min/week exhibited a lower incidence of AS (HR, 0.82; 95% CI, 0.73-0.91) compared to individuals with 0 min/week of MVPA. In contrast, despite the HR trends suggesting a potential decrease in the risk of AR and MR with MVPA volume, no statistically significant associations were observed (Figure 4).

**Figure 3.**
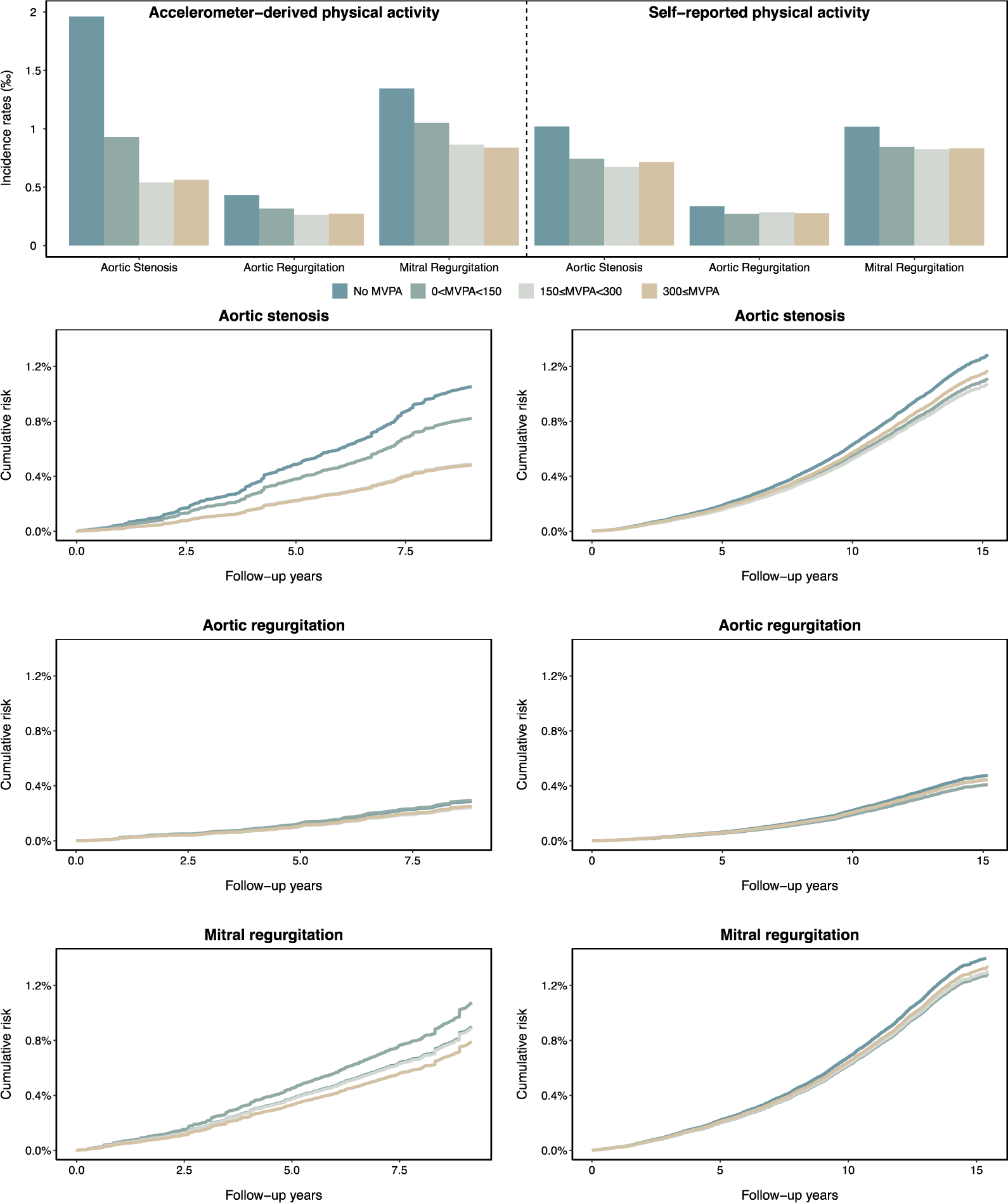
Incidence rates and adjusted survival curves for valvular heart disease by moderate-to-vigorous intensity physical activity volume groups. Upper panel. Incidence rates per 1,000 person-years of left-sided degenerative valvular heart disease by moderate-to-vigorous intensity physical activity volume groups. Lower panel. The left column represents adjusted survival curves for valvular heart disease incidence by accelerometer-measured physical activity groups. The right column represents adjusted survival curves for valvular heart disease incidence by questionnaire-based physical activity groups. Adjusted for age, sex, ethnicity, Townsend deprivation index, educational attainment, smoking status, alcohol consumption, diet score, sleep score, discretionary screen-time, and clinical comorbidities, including hypertension, obesity, type 2 diabetes, dyslipidemia, ischemic heart disease, stroke, cardiomyopathy, atrial fibrillation, chronic obstructive pulmonary disease, end-stage renal disease, and cancer. MVPA, moderate-to-vigorous intensity physical activity.

**Figure 4.**
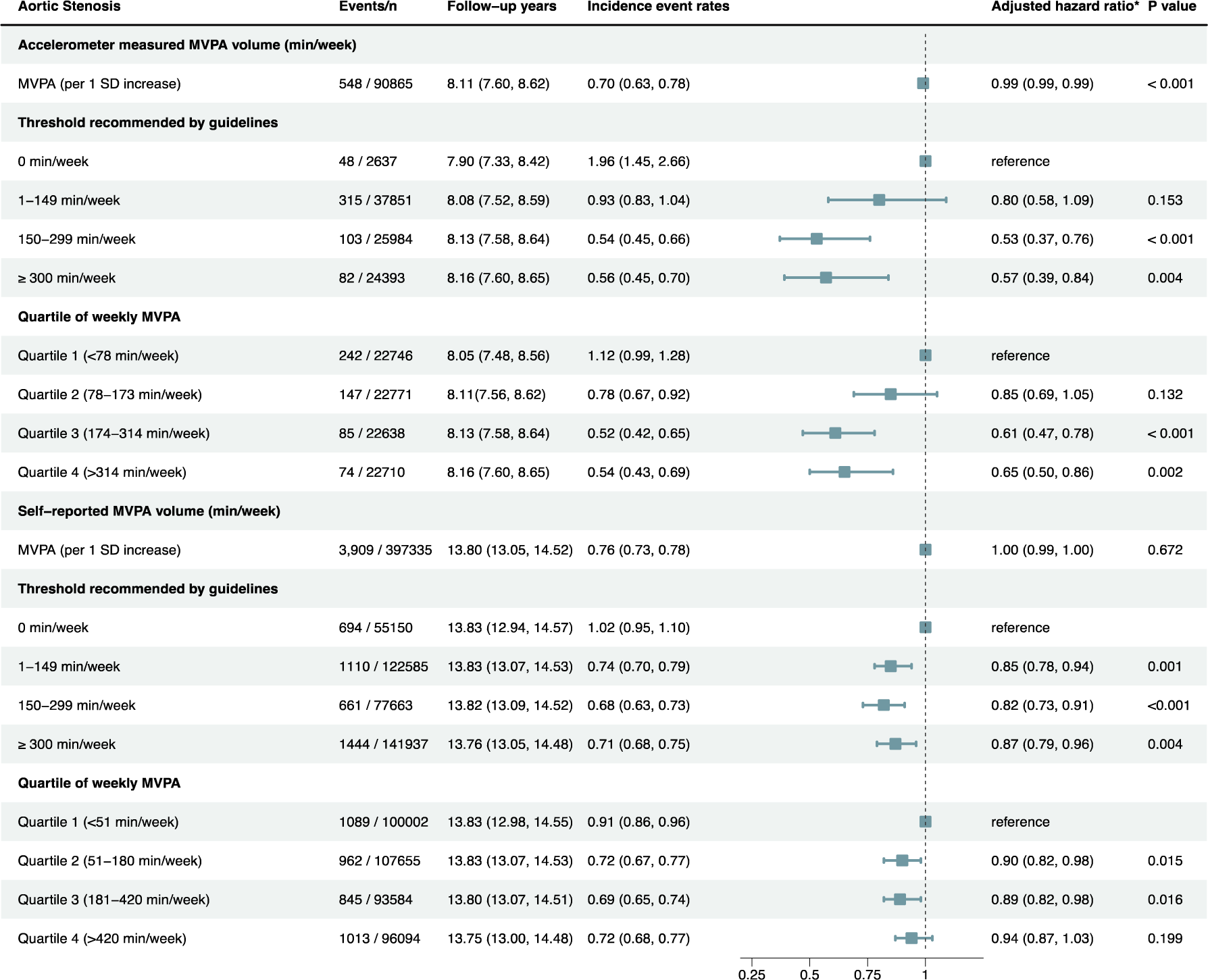
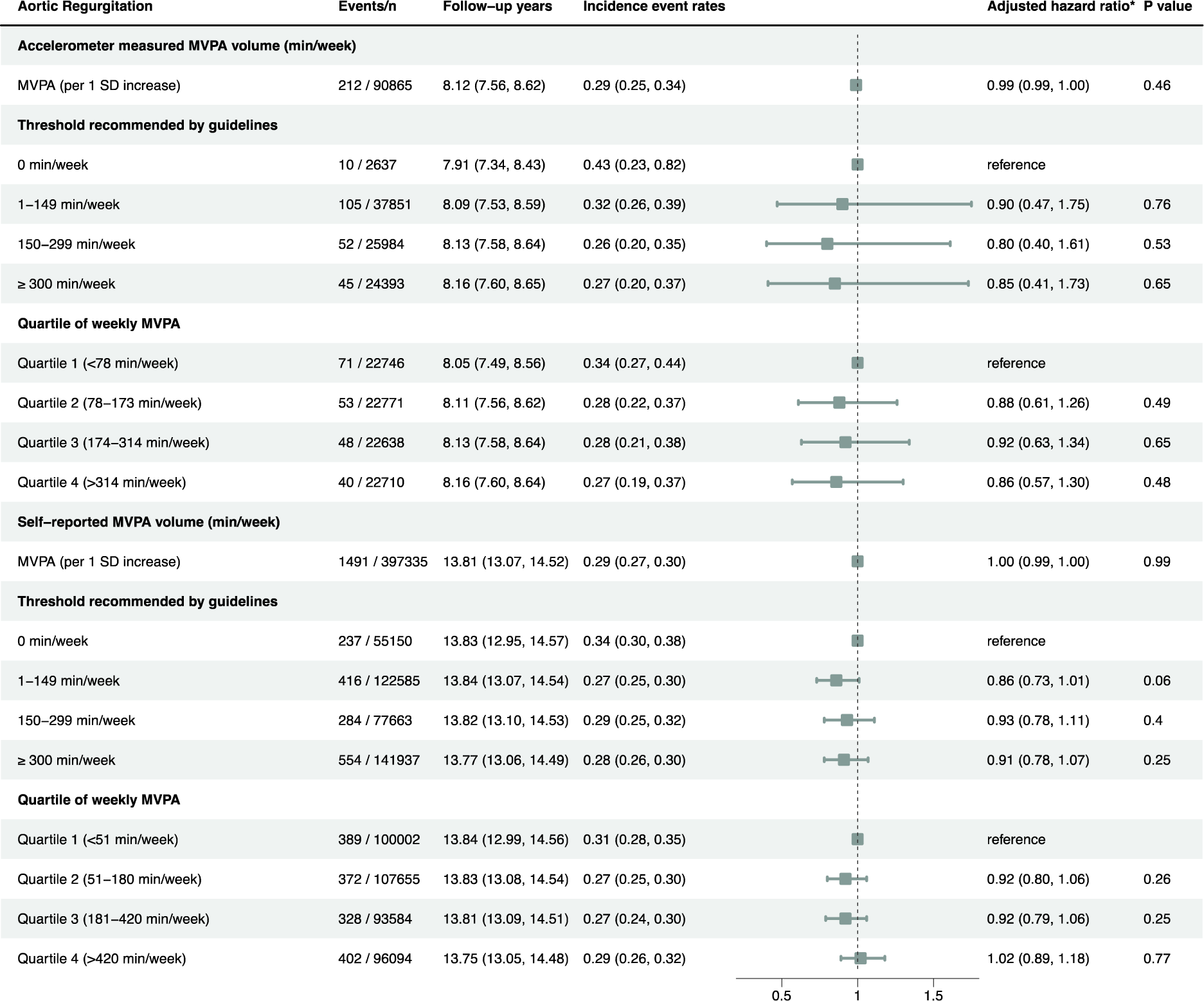
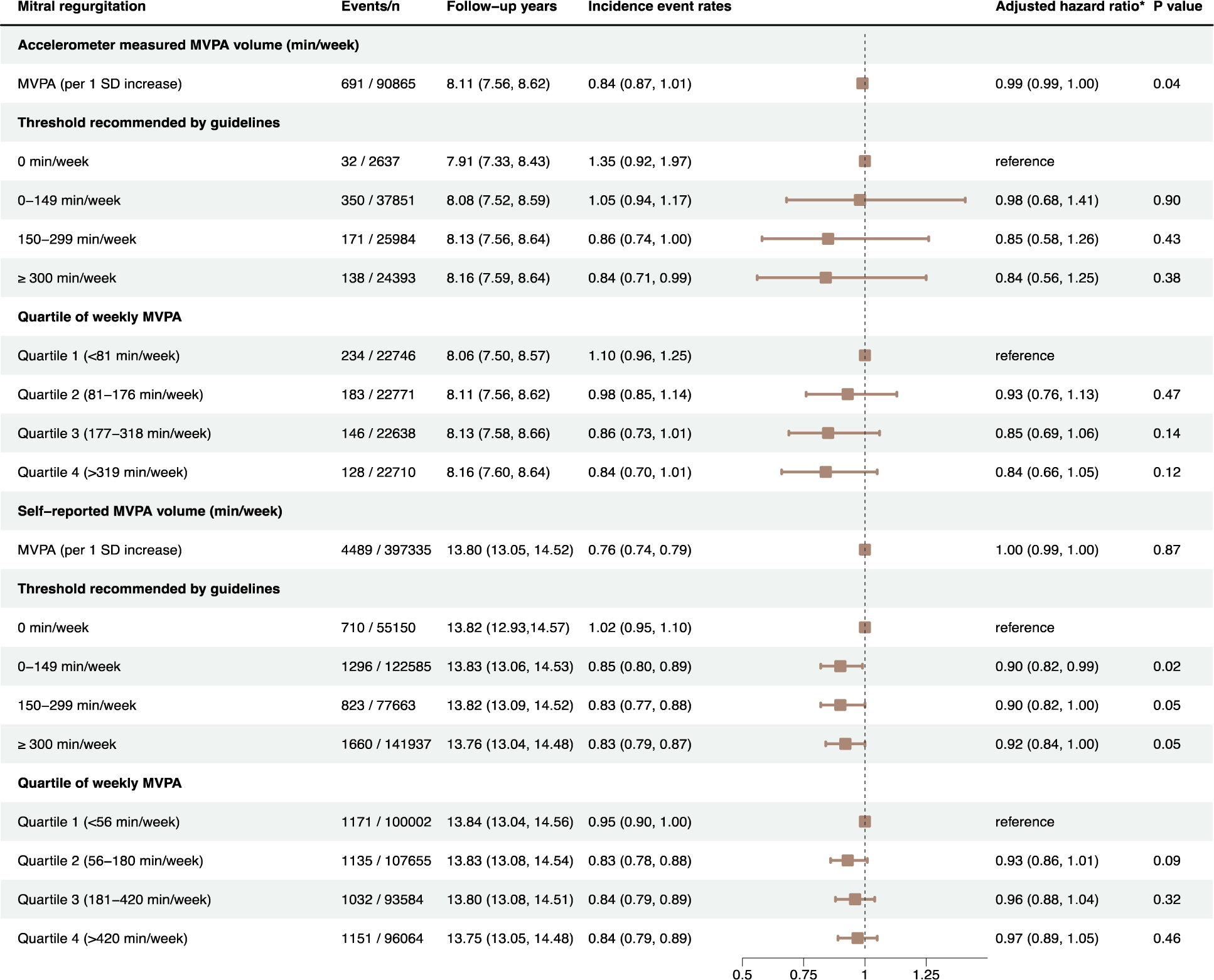
The association between the moderate-to-vigorous intensity physical activity and incident left-sided degenerative valvular heart disease. Incident event rates of left-sided degenerative VHD within MVPA volume groups were adjusted for age and gender and reported as events per 1,000 person-years using Poisson regression.*Adjusted for age, sex, ethnicity, Townsend deprivation index, educational attainment, smoking status, alcohol consumption, diet score, sleep score, discretionary screen-time, and clinical comorbidities, including hypertension, obesity, type 2 diabetes, dyslipidemia, ischemic heart disease, stroke, cardiomyopathy, atrial fibrillation, chronic obstructive pulmonary disease, end-stage renal disease, and cancer. MVPA, moderate-to-vigorous intensity physical activity.

### Consistent inverse association of MVPA volume with AS incidence

The main results remained robust across various sensitivity analyses. Accounting for competing risks from other causes of death (Figure S6-S8) and excluding participants who had a VHD event within 2 years of follow-up (Figure S9-S11) led to slightly attenuated associations. Analyzing the complete case sample, which included 88,210 individuals in the primary cohort and 385,699 individuals in the secondary cohort, consistently demonstrated the inverse association between guideline-adherent MVPA volume and AS incidence (Figure S12-14). Furthermore, additional analyses adjusting for systolic blood pressure, body mass index, lipoprotein (a), low-density lipoprotein cholesterol, glycated hemoglobin A1c and estimated glomerular filtration rate as covariates did not significantly alter the associations identified in the main analyses (Figure S15-17).

The inverse association between MVPA volume and AS incidence remained consistent across different age groups (<65, ≥65), sex (male, female), Townsend deprivation index (<median, ≥median), hypertension (presence or absence), obesity (presence or absence), dyslipidemia (presence or absence), type 2 diabetes (presence or absence), and cancer (presence or absence), with all P for interaction>0.05 (Figure 5).

**Figure 5.**
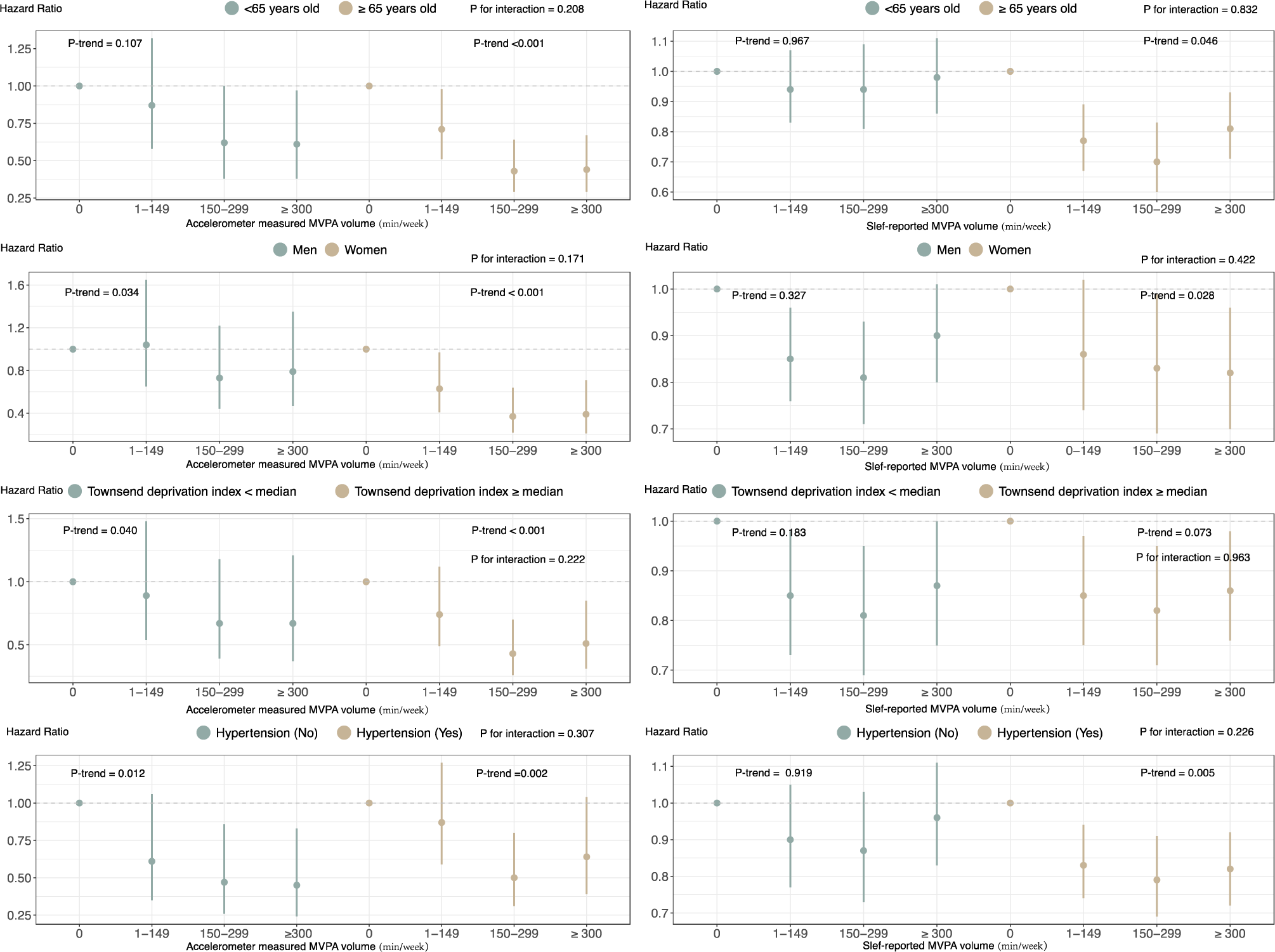
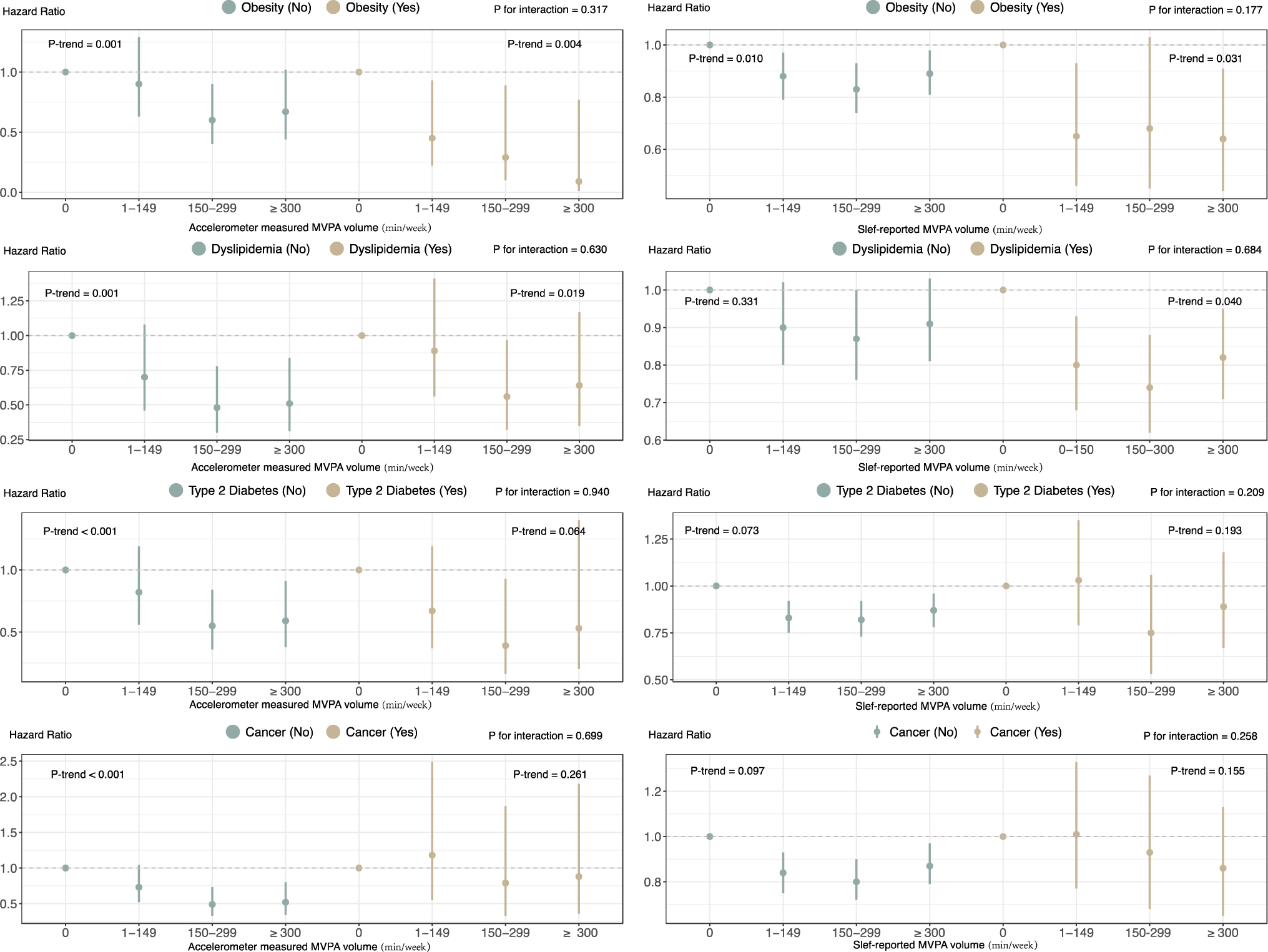
Subgroup analysis on the association between moderate-to-vigorous intensity physical activity volume groups and aortic valve stenosis incidence. Adjusted for age, sex, ethnicity, Townsend deprivation index, educational attainment, smoking status, alcohol consumption, diet score, sleep score, discretionary screen-time, and clinical comorbidities, including hypertension, obesity, type 2 diabetes, dyslipidemia, ischemic heart disease, stroke, cardiomyopathy, atrial fibrillation, chronic obstructive pulmonary disease, end-stage renal disease, and cancer. MVPA, moderate-to-vigorous intensity physical activity.

### Absolute 5-year and 10-year risk of AS by MVPA volume group

Higher MVPA levels and younger age are associated with lower 5-year and 10-year absolute AS risk, with female having lower risk than male (Figure 6). For instance, for individuals aged 65-74, the 5-year absolute risk of AS incidence was approximately three times higher among those with 0-149 min/week of accelerometer-measured MVPA compared to those with 150-300 min/week and ≥300 min/week of MVPA (Male: 10.25‰, 3.81‰, 4.78‰; Female: 5.67‰, 1.41‰, 2.09‰). The discrepancy in absolute risk was more pronounced in the accelerometer-measured physical activity cohort compared to the questionnaire-based physical activity cohort.

**Figure 6.**
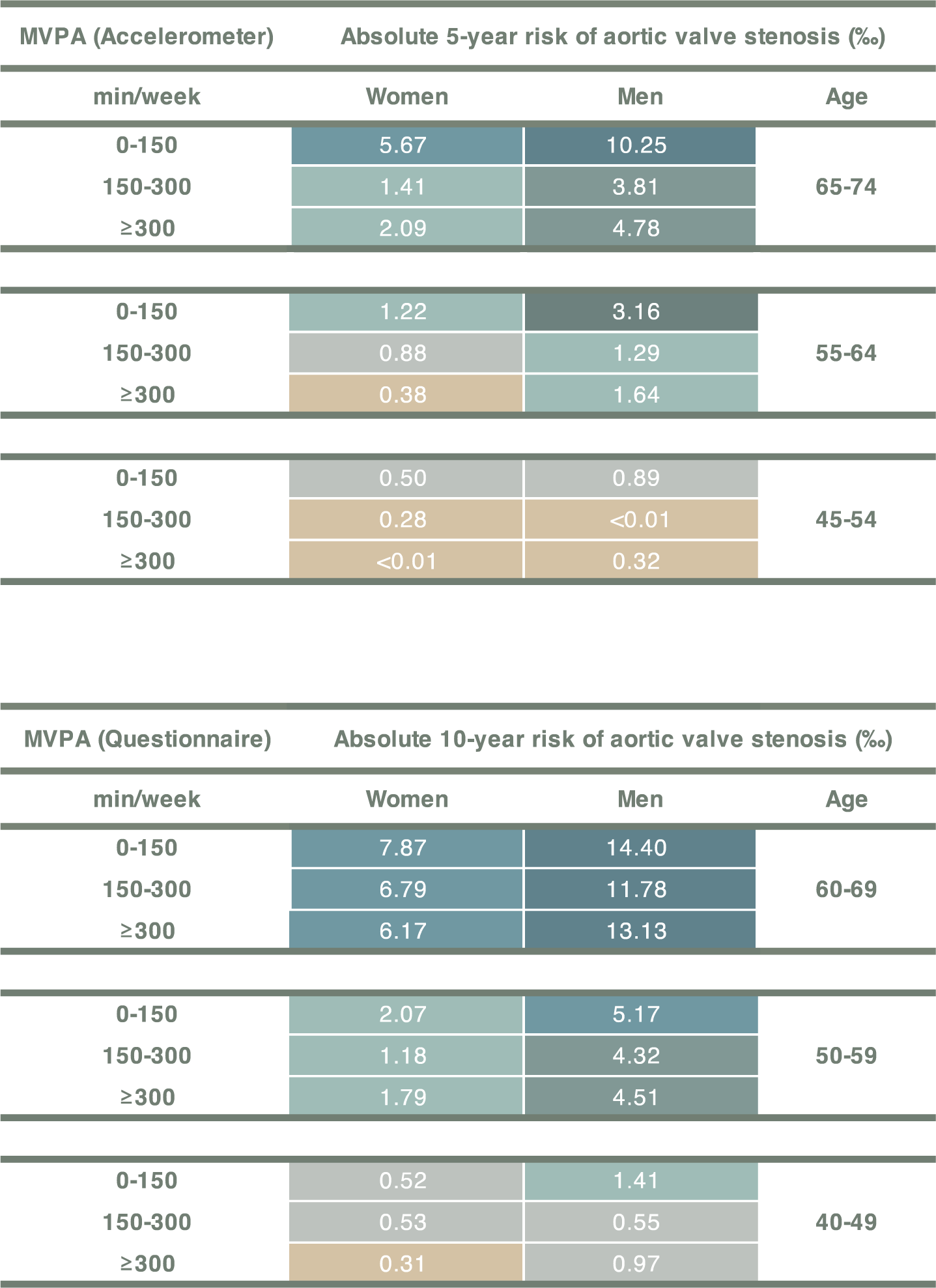
Absolute 5-year and 10-year risk of degenerative aortic valve stenosis. In the Fine and Gray’s competing risks regression model, we estimated the 5-year absolute risk of aortic valve stenosis in the primary cohort and the 10-year absolute risk in the secondary cohort. Age, sex, and physical activity categories were included as covariates, and multiple combinations of covariate values were employed to estimate the absolute risks. Numbers in grids are risk in per mille. MVPA, moderate-to-vigorous intensity physical activity.

### Association of MPA volume, VPA volume with AS incidence

In the exploratory analysis, we observed a significant association between MPA and a lower incidence of AS, while no significant association was found for VPA (Figure S18-19). Moreover, when evaluating individuals with an equivalent volume of MVPA, a higher ratio of VPA to total MVPA was not associated with a lower incidence of AS (Figure S19). These findings suggest that engaging in MPA within the current recommended range (150 to 299 minutes per week) may be more beneficial for preventing AS. The joint association analysis of MPA and VPA demonstrated that participants engaging in 150 to 299 minutes per week of MPA and 1 to 74 minutes per week of VPA exhibited the lowest risk of AS within the accelerometer-measured physical activity cohort (HR, 0.53; 95% CI, 0.37-0.76), as detailed in Table 2, Table S8, and Table S9.

**Table 2.** Adjusted hazard ratios for joint associations of MPA volume and VPA volume with aortic valve stenosis incidence.

|  |  | MPA, hazard ratio (95% CI) |  |  |
| --- | --- | --- | --- | --- |
| Accelerometer |  | 1-149 min/week | 150-299min/week | ≥300 min/week |
|  | 1-74 min/week | 0.76 (0.55, 1.05) | 0.53 (0.37, 0.76) * | 0.56 (0.38, 0.83) * |
| VPA | 75-149 min/week | - | 1.00 (0.38, 2.82) | 0.64 (0.25, 1.63) |
|  | ≥150 min/week | - | - | 0.99 (0.30, 3.21) |

|  |  | MPA, hazard ratio (95% CI) |  |  |  |
| --- | --- | --- | --- | --- | --- |
| Questionnaire |  | 0 min/week | 1-149 min/week | 150-299min/week | ≥300 min/week |
|  | 0 min/week | 1.00 (reference) | 0.90 (0.81, 1.00) | 0.87 (0.75, 1.02) | 0.90 (0.79, 1.03) |
| VPA | 1-74 min/week | 0.94 (0.68, 1.30) | 0.72 (0.63, 0.82) * | 0.81 (0.69, 0.96) | 0.75 (0.65, 0.88) * |
|  | 75-149 min/week | 1.13 (0.75, 1.71) | 0.83 (0.70, 0.99) | 0.81 (0.66, 1.00) | 0.95 (0.81, 1.12) |
|  | ≥150 min/week | 1.00 (0.67, 1.49) | 0.81 (0.64, 1.01) | 0.86 (0.71, 1.03) | 0.90 (0.79, 1.02) |

\* P values remained significant after multiple testing with the FDR methods. The reference group for this analysis consisted of individuals who had the lowest levels of both MPA (0 min/week) and VPA (0 min/week). Adjusted for age, sex, ethnicity, Townsend deprivation index, educational attainment, smoking status, alcohol consumption, diet score, sleep score, discretionary screen-time, and clinical comorbidities, including hypertension, obesity, type 2 diabetes, dyslipidemia, ischemic heart disease, stroke, cardiomyopathy, atrial fibrillation, chronic obstructive pulmonary disease, end-stage renal disease, and cancer. The blanked cells contained an insufficient number of participants to allow for the estimation of hazard ratios.

## Discussion

In this large-scale prospective study of middle-aged individuals, we identified a robust non-linear inverse association between MVPA volume and the risk of AS. Those who adhered to the recommended 150-299 minutes of accelerometer-measured MVPA per week showed a significant 50% reduction in AS risk compared to those reporting no MVPA. The beneficial association was consistently observed in high-risk populations for AS, including the elderly and individuals with hypertension, obesity, dyslipidemia, and type 2 diabetes, suggesting that targeted physical activity during the early stages of the disease may contribute to reducing the incidence of AS. Further investigations indicated that individuals with a higher proportion of VPA did not experience additional benefits for the same overall MVPA volume, highlighting the potential benefits of moderate-intensity activity in preventing AS. However, we did not find a consistent inverse association between MVPA volume and the incidence of AR and MR, indicating distinct pathological mechanisms underlying the development of valvular stenotic lesions and valvular regurgitation lesions.

### Distinct modalities of physical activity assessment on AS Risk

The existing epidemiological evidence on the association between physical activity and VHD incidence is limited. To the best of our knowledge, only two studies have investigated the association with AS risk. Eveborn et al. found no associations between questionnaire-based physical activity and intensity with AS risk.^10^ Similarly, Sarajlic et al. reported no associations between self-reported activities such as ≥1 hour/day of walking or ≥4 hours/week of exercise and the incidence of AS.^9^ These findings should be interpreted with caution as they are limited by non-standardized activity questionnaire content and unclear AS endpoints. Recent studies have indicated that activity levels measured using accelerometers may serve as a more reliable predictor of cardiovascular outcomes in comparison to questionnaire-based assessments within the same population.^21, 22^ Our observations align with previous studies, demonstrating significantly stronger effect sizes and improved risk stratification when employing accelerometer-derived measures of physical activity compared to self-reported assessments. Engaging in 150-299 minutes of accelerometer-measured MVPA per week reduces AS risk by nearly 50%, while questionnaire-based MVPA of the same duration lowers the risk by around 20%.

### Various intensity of physical activity on AS Risk

Our findings generally support the idea of achieving guideline thresholds (150 min/week) for MVPA to reduce the risk of AS. These associations were observed up to approximately 150-299 minutes per week, with MPA showing the most benefits. However, no significant association was found between accelerometer-measured VPA, the percentage of VPA in total physical activity, and the risk of AS. The limited number of individuals meeting the guideline-recommended VPA threshold of 75 minutes per week, coupled with the scarcity of events at longer VPA durations or higher percentages of VPA in total physical activity, could potentially obscure the true dose-response relationship. This observation is in line with previous studies that employed different quantification methods for VPA using wrist-worn accelerometers. ^20, 27^ It could imply a relatively lower engagement in vigorous physical activity among the middle-aged population, possibly due to the heightened challenges associated with vigorous-intensity activity. Subsequent research should delve into the potential impact of prolonged VPA on the risk of AS.

### Underlying Mechanism of Physical Activity Impact on VHD

Our findings are in line with recent laboratory studies that show the preventive effects of regular physical activity on aortic valve sclerosis in mice.^35^ This protection is attributed to maintaining valvular endothelial integrity, reducing inflammation and oxidative stress, and inhibiting the osteogenic pathway.^35^ Additionally, extensive research has revealed several protective associations linked to physical activity, such as improved insulin sensitivity, enhanced lipid metabolism, increased blood flow perfusion, and accelerated adipose metabolism.^36, 37^ These findings are important given the role of dyslipidemia, local inflammation, and endothelial injury as key factors in AS development.^38, 39^ Mechanisms of valvular regurgitation lesions may differ from valvular stenotic lesions. Evidence from mendelian randomization and observational studies does not support the notion that cardiometabolic risk factors for AS, such as low-density lipoprotein cholesterol and type 2 diabetes, are modifiable risk factors for AR and MR.^30, 40^ These findings align with the understanding that degenerative MR, primarily caused by mitral valve prolapse, is a common heritable valvulopathy characterized by fibromyxomatous changes in the mitral leaflet tissue. ^41^ Hence, these factors may contribute to the observed differences in the effects of physical activity on various types of VHD outcomes.

### Clinical implications

Given the current lack of effective preventive strategies for AS, our study provides novel insights into the potential role of increasing MVPA levels to reduce AS burden. Our research further elucidates the potential thresholds of benefit in relation to the volume and intensity of physical activity. Adhering to a moderate volume (150-299 minutes per week) and moderate intensity of physical activity (e.g., walking at a speed of 2.5 miles per hour) may be more feasible for middle-aged individuals at higher risk for AS. This approach takes into account the physical limitations associated with lower fitness levels in this age group, where achieving longer durations and higher intensities of physical activity could be more challenging.^42, 43^

With the rising popularity of smart wearable devices, monitoring physical activity has become more accessible.^44^ The utilization of wearable devices allows individuals to receive real-time feedback on their weekly activity levels. This technological advancement provides valuable and actionable information for personalized prevention strategies.^45^ Our findings suggest that adhering to activity thresholds measured through wearable devices may enhance the effectiveness of reducing AS risk when compared to using subjective physical activity questionnaires. These findings have the potential to inform future VHD guideline recommendations, aiming to alleviate the healthcare burdens associated with AS. Furthermore, recent clinical trials have highlighted the significant role of physical activity in both primary and secondary cardiovascular prevention.^7, 8^ Building upon these findings, our study lays a foundation for future clinical trials aimed at exploring the potential of MVPA as a secondary prevention strategy to slow the progression of AS.

### Limitations

Several factors should be considered when interpreting our findings. Firstly, while wearable devices offer more precise measurements compared to questionnaire-based assessments of physical activity, they may not fully capture individuals engaged in non-ambulatory activities such as resistance exercise or indoor cycling. Future research should explore the impact of different types of physical activity on AS risk. Secondly, relying solely on ICD-10 diagnosis codes may mainly capture advanced stages of VHD, potentially leading to an underestimation of disease risk due to the underdiagnosis and late identification of milder, asymptomatic cases. Moreover, this approach does not provide information on the severity of valvular stenosis or regurgitation, which presents challenges for further analysis on the effectiveness of physical activity as a secondary prevention strategy. Additionally, specific subgroup analyses, such as individuals with end-stage renal disease, may have limited statistical power due to lower event rates. Thirdly, the main UK Biobank sample is not representative of the overall UK population, as it consists predominantly of individuals of White European ancestry who generally have better health compared to the general population. However, it is important to note that while this sample may not accurately estimate the prevalence and incidence of health conditions, it does not impact the magnitude of the risk estimates. Previous research suggests that the exposure-outcome associations observed in the UK Biobank provide valid estimates and align with results obtained from more representative samples.^46^ Besides, as an observational study, we cannot rule out the possibility of residual confounding, despite adjusting for a wide range of confounders in our analysis. Additionally, the presence of reverse causality should be considered, whereby individuals with VHD at the initial phase, often accompanied by other cardiovascular diseases, may have reduced physical activity. To address the issue, we conducted sensitivity analyses by excluding individuals who experienced an incident degenerative VHD event within the initial 2 years of follow-up, and the inverse associations persisted.

## Conclusions

Engaging in 150-299 min/week of MVPA is associated with the lowest risk reduction for AS, with most of the benefits could be obtained at MPA. Objective activity monitoring through wearable devices shows promise as an effective nonpharmaceutical intervention to alleviate the healthcare burdens of AS.

## Contributors

All authors read and approved the final version of the manuscript. Z.L. proposed the idea, performed the data analyses and drafted the manuscript. P.P., M.C., S.C., L.D., Y.S., B.G., J.L., Y.H., T.L., X.G., checked the integrity and plausibility of data analysis. M.C., B.Z., H.X. and Y.W. revised the manuscript and was responsible for the integrity of data acquisition and statistical analyses. Z.L., B.Z., and Y.W. verified the underlying data.

## Data Availability

his study was conducted based on the UK Biobank cohort study under application number 91305.

